# Role of physiotherapy team in critically ill COVID-19 patients pronation: can a multidisciplinary management reduce the complications rate?

**DOI:** 10.1101/2021.06.20.21258949

**Authors:** Andrea Glotta, Nicola Faldarini, Maira Biggiogero, Andrea Saporito, Diana Olivieri, Claudia Molteni, Stefano Petazzi, Romano Mauri, Xavier Capdevila, Samuele Ceruti

## Abstract

**Objectives:** During the pandemic, critically ill COVID-19 patients’ management presented an increased workload for Intensive Care Unit (ICU) nursing staff, particularly during pronation maneuvers, with high risk of complications. In this scenario, a support during pronation by the *ICU Physiotherapy Team* was introduced.

**Research methodology:** Retrospective analysis. Consecutive critically ill COVID-19 patients.

**Setting:** A COVID-19 Center in southern Switzerland, between March 16^th^ and April 30^th^, 2020.

**Main Outcome Measures:** Rates and characteristics of pronation-related complications.

**Results:** Forty-two patients on mechanical ventilation (MV) were treated; 296 standard prone/supine positioning were performed, with 3.52 cycles/patient. One (0.3%) major complication was observed, while fourteen (33.3%) patients developed minor complications, e.g. pressure injuries. The incidence of pressure sores was related to ICU length-of-stay (LOS) (p = 0.029) and MV days (p = 0.015), while their number (n = 27) further correlated with ICU LOS (p = 0.001) and MV days (p = 0.001). The propensity matching score analysis did not show any protective factor of pronation regarding pressure injuries (p = 0.448). No other significant correlation was found.

**Conclusion:** The specific pronation team determined a low rate of major complications in critically ill COVID19 patients. The high rate of minor complications appeared to be related to disease severity, rather than from pronation.

## INTRODUCTION

COVID-19 pneumonia is a viral disease caused by SARS-CoV-2, affecting over 100,000,000 patients with 2,000,000 deaths worldwide (John Hopkins University and Medicine, 2020). One main aspect in critically ill COVID-19 patients concerns acute respiratory failure due to severe interstitial bilateral pneumonia, often requiring mechanical ventilation (MV) through oro-tracheal intubation (OTI) or tracheostomy if MV is prolonged (Alhazzani et al., 2020; Miles et al., 2020; Ranieri et al., 2012). In this scenario, the burden of COVID-19 patient management for the ICU nursing staff is greatly increased (Lucchini et al., 2020b). Pronation maneuvers are part of these patients management, especially in patients requiring mechanical ventilation for respiratory failure and Acute Respiratory Distress Syndrome (ARDS) [REF], in order to improve tissue oxygenation through an improvement in ventilation/perfusion mismatch (Apte et al., 2021; Jahani et al., 2018) this maneuver requires a degree of physical effort and the presence of multiple nurses for a correct execution of the technical gesture (Lucchini et al., 2018; Qadri et al., 2020).

Conflicting data regarding pronation-related complications in ARDS patients have been reported. Some authors described an increase in complications incidence after pronation (Kopterides et al., 2009; Sud et al., 2014, 2010; Taccone et al., 2009), while other groups did not find an increased risk due to pronation (Abroug et al., 2011; Guérin et al., 2018, 2013), perhaps because of the presence of highly experienced and qualified personnel (Abroug et al., 2011; Guérin et al., 2013; Lucchini et al., 2020a; Simonelli et al., 2020). In critically ill COVID-19 patients, some groups addressed the problem of high risk of complication related to this increased amount of pronation (Cotton et al., 2020; Kimmoun et al., 2020). In this scenario, B. Short et al (Short et al., 2020) reported a clinical benefits in having a specific “pronation team” dedicated to patients’ pronation during the pandemic wave (Doussot et al., 2020), even though they focused mainly over clinical patients evolution, rather than over the rate of complications related to this delicate procedure.

During the first weeks of the pandemic, a high ICU workload was immediately registered (Lucchini et al., 2020b), including an increased number of pronations, with consequent increase in related complications incidence. In this emergency scenario, our Clinic, having been identified as a COVID-19 Center by the Swiss Health Department [2], decided to introduce a dedicated *ICU Physiotherapy Team* (IPT) (Simonelli et al., 2020) to support nursing staff in the implementation of a correctly executed pronation maneuvers. The aim of this approach was to reduce the risk of complications through the use of qualified personnel dedicated to patient mobilization; the IPT was provided therefore a daily pronation treatment for critically ill patients with COVID-19 (Vitacca et al., 2020).

## METHODS

We performed a retrospective analysis on consecutive critically ill COVID-19 patients; those untreated by the IPT were excluded. The objective of this study was to retrospectively analyze complications rates related to pronation in this ongoing process of collaboration between physiotherapists and nurses in critically ill COVID-19 patients, evaluating their interaction with outcomes on duration of MV and ICU length-of-stay (LOS). Demographic data such as age and Body-Mass Index (BMI), comorbidities such as chronic-obstructive pulmonary disease (COPD), obstructive sleep apnea syndrome (OSAS), diabetes, hypertension and ischemic heart disease (IHD) were registered and reported. Evaluation of ICU severity scores at admission (NEMS - Nine equivalents of nursing manpower use score -, SAPS - Simplified Acute Physiology Score - and SOFA - Sequential Organ Failure Assessment -), and MV features such as OTI, need for tracheostomy, days of MV, ICU LOS and number of pronation maneuvers were also reported. Ventilated patients were further stratified according to the MV modality, subdividing invasive MV (OTI and tracheostomy) and non-invasive MV (C-PAP, High-Flow or nasal cannula oxygen-therapy). Data regarding actual/past ICU workload (NEMS, SAPS and SOFA) were extrapolated from a public national dataset (“Registro dei dati - SGI-SSMI-SSMI Società svizzera di medicina intensiva,” n.d.) through queries concerning *critical patients [admitted in ICU] AND first pandemic wave [from March 16*^*th*^ *2019 to April 16*^*th*^ *2019] AND [primary diagnosis of pulmonary infection]*; all data were collected and reported in the electronic medical record.

### Indication to pronation

Criteria for pronation included critically ill COVID19 patients on invasive MV with ARDS criteria (Ranieri et al., 2012) and a P/F-ratio less than 150 despite standard-of-care management, associated with a protective MV (low-volume, low-pressure ventilation and adequate ventilator synchrony) (Brower et al., 2004) with FiO_2_ more than 60% and PEEP titrated according to ARDS network (Brower et al., 2004). Pronation cycles were continued until the P/F-ratio remained less than 150 when supine, with FiO_2_ greater than 60% and PEEP greater than 10 cmH_2_O.

### ICU Physiotherapy Team

The IPT consisted of a nurse and a minimum of 4 physiotherapists, of which one was specialized physiotherapist in cardiorespiratory and critical care rehabilitation. Activities in the ICU were organized with a daily employment of 4/5 physiotherapists for 7 days per week. Based on the patient clinical situation and evolution, after a short assessment with the Intensivist in charge, pronations took place every day at 4.00 pm; the IPT provided assistance to the nursing staff for critically ill COVID-19 patients’ systematic pronation. The supinations occurred at 8.00 am, if clinicians did not provide a different indication. Average length of pronation was 16 hours a day.

The pronation gesture was executed according to international standard (Doussot et al., 2020; Wiggermann et al., 2020). During the pronation maneuvers, the nurse in charge managed the head and the main devices (endotracheal tube, central venous catheter, dialysis catheter); the nurse in charge was also responsible for the IPT coordination during the pronation and ensured maneuver correct execution times. Physiotherapists were positioned in pairs on both sides of the patient, to perform pronation according to the nurse’s indications. Two physiotherapists managed pronation of the thoraco-abdominal area, while other two managed the legs and the urinary catheter. Data concerning total number of patient pronation cycles and all pronations-related data were recorded in the electronic medical database.

### Complications

According to other groups (Kopterides et al., 2009; McCormick and Blackwood, 2001; Park et al., 2015; Sud et al., 2014), adverse events pronation-related were stratified into major and minor complications. *Major complications* were intended as accidental endotracheal tube displacement, displacement of other devices (central venous catheter, thoracic or abdominal drainage), loss of peripheral vascular access, bone dislocations. *Minor complications* were especially intended as the presence of pressure ulcers, reported according to severity stage, number of sores and location (Kopterides et al., 2009; McCormick and Blackwood, 2001; Park et al., 2015; Sud et al., 2014). All information reported in the electronic medical record has been collected.

### Outcomes

The primary outcome was to retrospectively analyze and report major complications rates in critically ill COVID-19 patients managed by the specialized team with standard cycles of pronation. Secondary outcomes were to analyze all correlations between the incidence of minor complications, like pressure injuries, and clinical data, so as to determine any potential protective factor.

### Statistical analysis

A descriptive statistic was conducted; data were reported as number (percentage). Data distribution was reported as mean (SD) or as median (IQR) according to the statistical distribution, verified by Kolmogorov-Smirnov test. Differences between patient outcomes were studied by t-test for independent groups or by Wilcoxon test if non-parametric analysis was required. Study of differences between groups of categorical data was carried out by Chi-square statistics. To assess the impact of treatment (pronation/non-pronation) on complications incidence and to control for confounding factors, a propensity score-adjusted multivariable analysis through available data was performed. Quantitative variables like age, BMI, MV days, ICU LOS, and the scores at admission SAPS, NEMS, SOFA were used; qualitative variables like sex, presence of tracheostomy or IOT, presence of comorbidities like hypertension, OSAS, COPD, diabetes, ischemic heart disease and presence of complications like VAP, need of CVVHDF during recovery, other types of infection and pulmonary thromboembolism were determined. The aim was to identify if the treatment “pronation” could act as a protective factor in the avoidance of complications. The level of significance was established to be p < 0.05. Statistical data analysis was performed using SPSS (version 25.0; IBM Corp, Armonk, New York, USA).

### Ethical consideration

This study has been notified to the Ethics Committees (Comitato Etico Cantonale, Chairman Prof. Zanini CE_TI 3763), and it has been approved in agreement to the local Federal rules. Informed consent was obtained from patients involved in the analysis.

## RESULTS

From March 16^th^ to April 30^th^ 2020, 43 consecutive critically ill COVID-19 patients were admitted to ICU; one was excluded from the study because he was not treated by physiotherapists. Forty-two patients were instead treated by the IPT. The median age was 67.5 years (56.7 – 73 yrs.), 35 (83.3%) were male, with a mean BMI of 28.3 Kg/m^2^ (18.6 – 41.1, SD 5.1). Thirty-six patients (86%) were on MV, 66% ventilated by OTI and 19% by tracheostomy. The median ICU LOS for all patients was 11 days (8 – 18); median length of MV days was 8 days (5 – 13). At admission, mean SAPS score was 46.1 (13 – 94, SD 18), median SOFA score 7 (4 – 8.25) and median NEMS score was 34.5 (18 - 39) (Table 1). The actual mean NEMS value in patients admitted to ICU for COVID-19 pulmonary complication resulted 30.5 (9 – 42, SD 10.1); this was compared to the mean NEMS (22.6 (9 – 48, SD 8.2) of patients with similar clinic characteristic admitted to the ICU during the previous year, resulting in a significant augmentation of workload (p < 0.001).

**TABLE 1.** Demographics.

|  | Characteristics | Enrolled (n = 42) |
| --- | --- | --- |
| **Demographics** | Age | 67.5 (56.75 – 73) |
|  | Male | 35 (83.3) |
|  | BMI | 28 (18.6 – 41.1, SD 5.1) |
| **Comorbidities** | Hypertension | 20 (47.6%) |
|  | Diabetes | 14 (33.3%) |
|  | OSAS | 6 (14.3%) |
|  | COPD | 5 (11.9%) |
|  | Heart Ischemic Disease | 9 (21.4%) |
|  | VTE | 7 (16.7%) |
| **Severity score at ICU admission** | NEMS | 34.5 (18 – 39) |
|  | SAPS II | 46.1 (13 – 94, SD 18.22) |
|  | SOFA | 7 (4 – 8.25) |
| **MV parameters** | Patients on MV | 36 (86%) |
|  | • Endotracheal tube | 28 (66%) |
|  | • Tracheostomy | 8 (19%) |
|  | Pronation maneuvers | 3.52 (0 – 8, SD 2.48) |
|  | ICU LOS (n = 42) | 11 (6 – 18) |
|  | MV Days (n = 36) | 8 (5 – 13) |

*Demographic characteristics at ICU admission. Data are presented as means (min-max, SD) or medians (IQR) for non-normally distributed variables.*

### Primary outcome

During the study period, 296 technical gestures of pronation and supination (148 pronation cycles) were performed by the IPT on MV patients (Table 2); 36 patients (86%) of the forty-two included in the study were on MV and underwent pronation and supination maneuvers, with an average of 3.52 cycles per patient (1 - 8, SD 2.47). During pronation, a major complication was observed in one patient (0.3%), consisting in the accidental dislocation of the endotracheal tube. The tube dislocation was confirmed by emergency bronchoscopy and was associated with sudden peripheral desaturation, without any subsequent clinical consequence after rapid management. No other major complications were registered (Table 2).

**TABLE 2.** Major Complications pronation-related.

|  |  |
| --- | --- |
| **Number of pronation** | **296** |
| **Endotracheal tube removal** | 0 |
| **Endotracheal tube dislocation** | 1 (0.3) |
| **Displacement of devices** | 0 |
| **Loss of peripheral vascular access** | 0 |
| **Bone dislocations** | 0 |

*Major complications in ICU directly related to the technical gesture of pronation. Data are presented as numbers and percentual.*

### Secondary outcomes

Despite the prone position, 14 (33.3%) patients presented minor complications consisting in pressure injuries; 8 (19%) patients showed a single pressure sore, 2 (4.8%) patients presented 2 lesions, 3 (7.1%) patients had 3 lesions and 1 (2.4%) patient presented 5 different pressure injuries. Data regarding lesion distribution and staging are reported in Table 3. The pressure injury distribution however did not involve areas with major pressure, with sacral decubitus occurring in 33.3% of patients, even if they remained pronated for 16 hours/day. The *number* of pressure injuries presented significant correlations with MV days (r = 0.475, p = 0.001) (Figure 1), and ICU LOS (r = 0.467, p = 0.001) (Figure 2); similarly, the *presence* of pressure injuries also correlated with ICU LOS (p = 0.029) and MV days (p = 0.015) (Figure 2). No significant correlation was found with other variables (Table 4).

**TABLE 3.** Stratification of minor complications pronation-related (pressures injuries) according degree and site.

|  | **Degree I** | **Degree II** | **Degree III** | **Degree IV** | **Total** |
| --- | --- | --- | --- | --- | --- |
| **Chest** | 1 (3.7) | 5 (18.5) | 0 | 0 | **6 (22.2)** |
| **Sacred** | 2 (7.4) | 3 (11.1) | 4 (14.8) | 0 | **9 (33.3)** |
| **Chin** | 0 | 2 (7.4) | 1 (3.7) | 0 | **3 (11.1)** |
| **Lip rhyme** | 0 | 1 (3.7) | 0 | 0 | **1 (6.25)** |
| **Knee** | 2 (7.4) | 1 (3.7) | 0 | 0 | **3 (11.1)** |
| **Cheek** | 0 | 2 (7.4) | 1 (3.7) | 0 | **3 (11.1)** |
| **Hip** | 0 | 1 (3.7) | 0 | 0 | **1 (6.25)** |
| **Gluteus** | 0 | 1 (3.7) | 0 | 0 | **1 (6.25)** |
| **Total** | **5 (18.5)** | **16 (59.2)** | **6 (22.2)** | **0** | **27 (100)** |

*Pressure injuries degrees and sites, intended as “minor complications” of pronation. The greatest prevalence of lesions occurs at sacral level, an anatomical area that mechanically should be preserved from compression with pronation. Data are presented as numbers and percentual.*

**TABLE 4:**
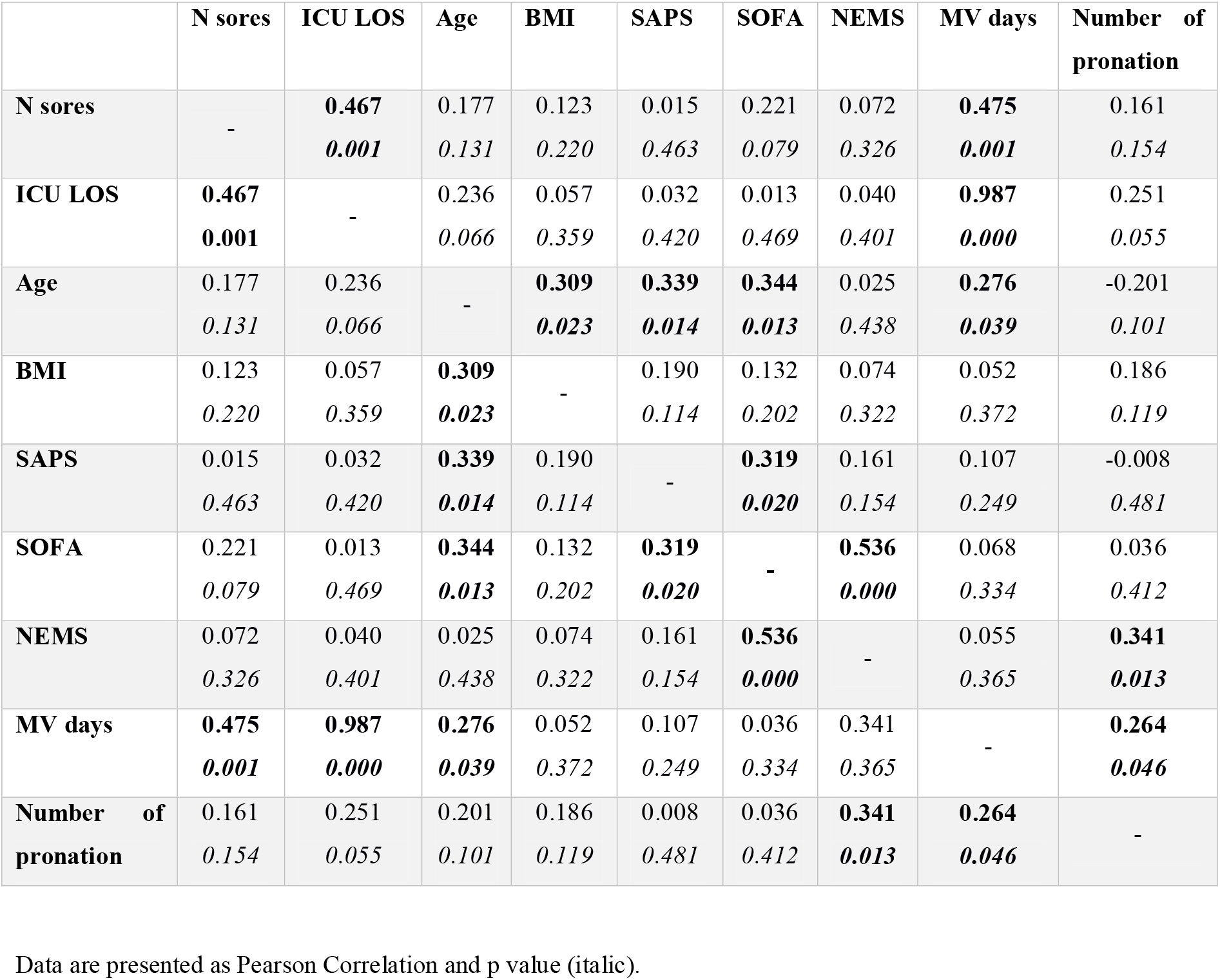
Correlation analysis between main variables.

|  | N sores | ICU LOS | Age | BMI | SAPS | SOFA | NEMS | MV days | Number of pronation |
| --- | --- | --- | --- | --- | --- | --- | --- | --- | --- |
| **N sores** | - | **0.467**<br>*0.001* | 0.177<br>*0.131* | 0.123<br>*0.220* | 0.015<br>*0.463* | 0.221<br>*0.079* | 0.072<br>*0.326* | **0.475**<br>*0.001* | 0.161<br>*0.154* |
| **ICU LOS** | **0.467**<br>*0.001* | - | 0.236<br>*0.066* | 0.057<br>*0.359* | 0.032<br>*0.420* | 0.013<br>*0.469* | 0.040<br>*0.401* | **0.987**<br>*0.000* | 0.251<br>*0.055* |
| **Age** | 0.177<br>*0.131* | 0.236<br>*0.066* | - | **0.309**<br>*0.023* | **0.339**<br>*0.014* | **0.344**<br>*0.013* | 0.025<br>*0.438* | **0.276**<br>*0.039* | -0.201<br>*0.101* |
| **BMI** | 0.123<br>*0.220* | 0.057<br>*0.359* | **0.309**<br>*0.023* | - | 0.190<br>*0.114* | 0.132<br>*0.202* | 0.074<br>*0.322* | 0.052<br>*0.372* | 0.186<br>*0.119* |
| **SAPS** | 0.015<br>*0.463* | 0.032<br>*0.420* | **0.339**<br>*0.014* | 0.190<br>*0.114* | - | **0.319**<br>*0.020* | 0.161<br>*0.154* | 0.107<br>*0.249* | -0.008<br>*0.481* |
| **SOFA** | 0.221<br>*0.079* | 0.013<br>*0.469* | **0.344**<br>*0.013* | 0.132<br>*0.202* | **0.319**<br>*0.020* | - | **0.536**<br>*0.000* | 0.068<br>*0.334* | 0.036<br>*0.412* |
| **NEMS** | 0.072<br>*0.326* | 0.040<br>*0.401* | 0.025<br>*0.438* | 0.074<br>*0.322* | 0.161<br>*0.154* | **0.536**<br>*0.000* | - | 0.055<br>*0.365* | **0.341**<br>*0.013* |
| **MV days** | **0.475**<br>*0.001* | **0.987**<br>*0.000* | **0.276**<br>*0.039* | 0.052<br>*0.372* | 0.107<br>*0.249* | 0.036<br>*0.334* | 0.341<br>*0.365* | - | **0.264**<br>*0.046* |
| **Number of pronation** | 0.161<br>*0.154* | 0.251<br>*0.055* | 0.201<br>*0.101* | 0.186<br>*0.119* | 0.008<br>*0.481* | 0.036<br>*0.412* | **0.341**<br>*0.013* | **0.264**<br>*0.046* | - |

Data are presented as Pearson Correlation and p value (italic).

**FIGURE 1:**
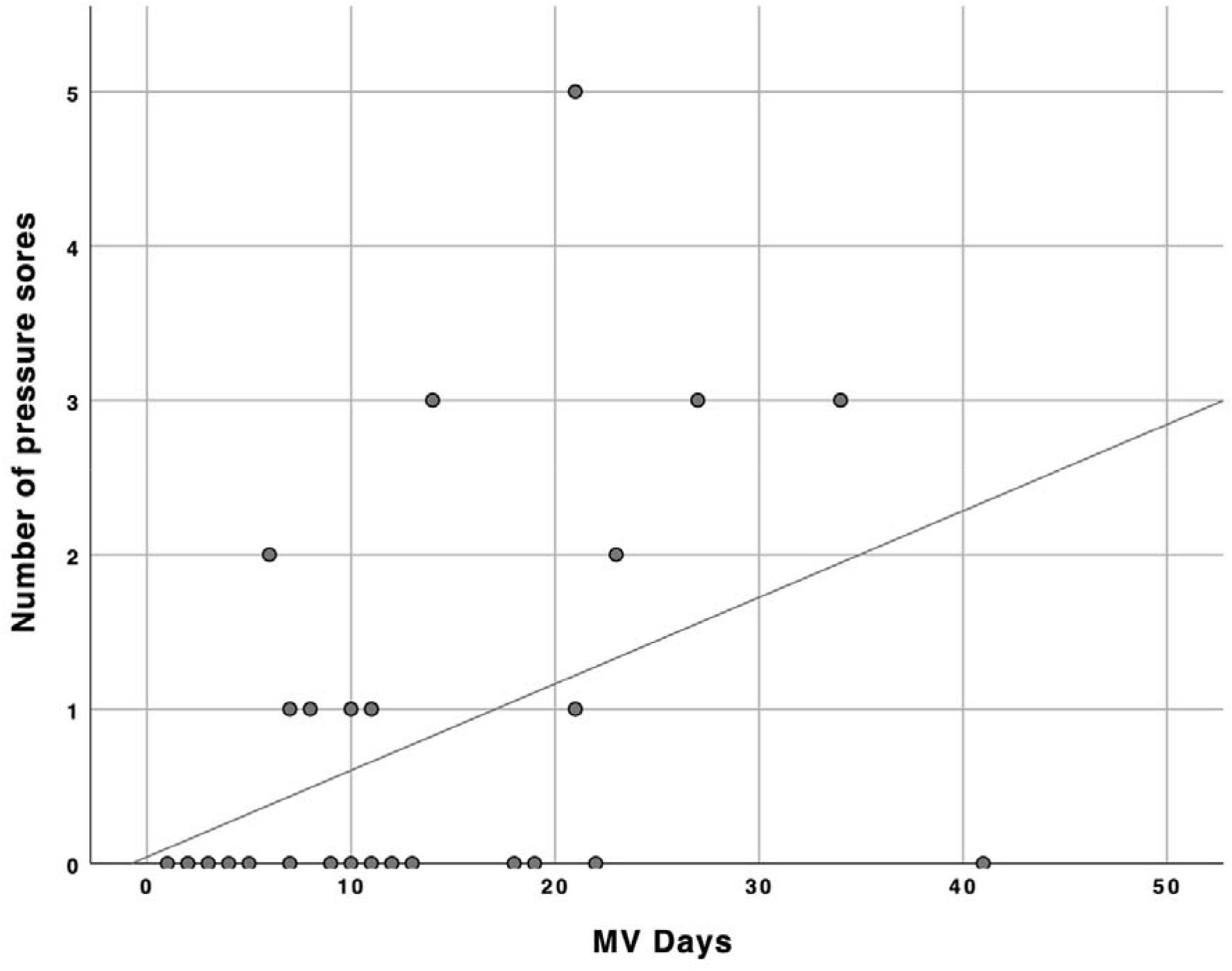
Patients’ distribution comparing number of MV days and pressure injuries (r = 0.475, p = 0.001).

**FIGURE 2:**
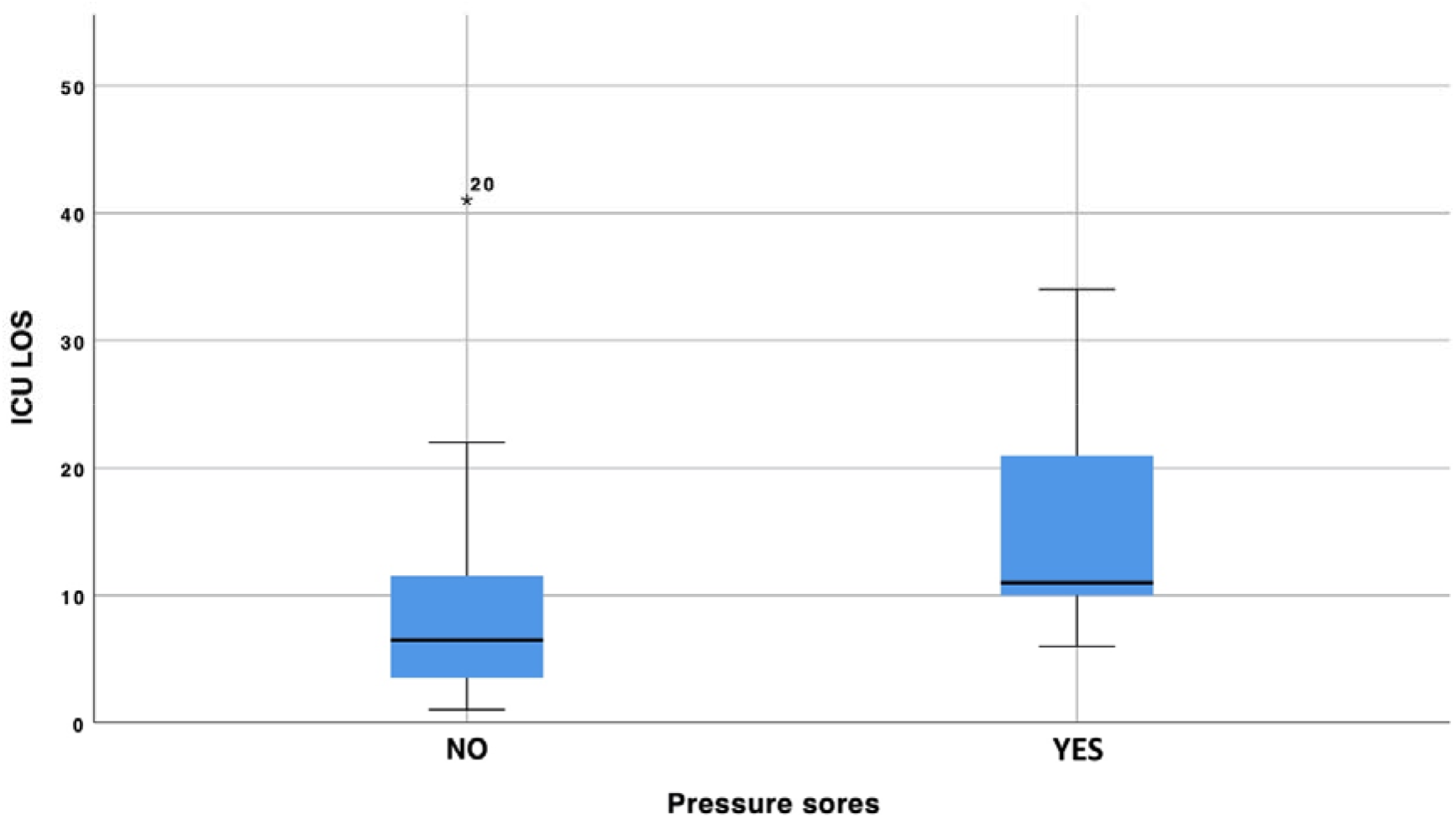
Distribution of ICU LOS correlated to the presence/absence of pressure injuries (p=0.029).

Because of the retrospective design of the study and the imbalance between treatment, especially regarding pronation/non pronation and the total number of pronations, propensity score was applied with the aim to better estimate the effect of observed data. The propensity matching score showed no protective factor of pronation regarding the incidence of pressure sores (p = 0.448).

## DISCUSSION

To date, several groups have approached the problem regarding the creation of a dedicated team in order to better manage the high number of pronations in critically ill COVID-19 patients (Cotton et al., 2020; Doussot et al., 2020; Johnson et al., 2021; Ng et al., 2020; Short et al., 2020). However, no studies reported data concerning the impact of a physiotherapy team on the incidence of pronation-induced complications. According to Lucchini et al [5], current data confirmed evidence that the nursing workload for critically ill COVID-19 patients was more increased compared to previous years. In this scenario, better procedural management require a multidisciplinary approach, with the aim to reduce pronation complication rates.

In the current study cohort, only one case of major complication induced by pronation was registered, and the presence of a specific IPT as a strategic aspect in the management of critically ill COVID-19 patients appeared to be a protective factor against the incidence of major complications. In accordance with Guerin et al. and Abroug et al. (Abroug et al., 2011; Guérin et al., 2013), we hypothesize therefore that a trained specialized team is essential to reduce major complications during pronation.

Different outcome resulted regarding minor complications. Current data induced to consider COVID-19 as a strongly debilitating disease in terms of catabolic energy balance (Romano et al., 2020). This hypothesis is supported by the identification of a direct correlation between MV days and ICU LOS with the number and the incidence of pressure sores, respectively. As already demonstrated by other group (Labeau et al., 2020; Strazzieri-Pulido et al., 2019), the greater the severity of the disease - also in catabolic terms - the greater the risk of pressure ulcers. Intriguingly, the extensive number of pressure injuries, even in anatomical district protected by pronation such as the sacral region, and their strong correlation with ICU LOS and MV days (intended as epiphenomena of disease severity), suggested that pronation did not play a protective effect on minor complications’ incidence. The pathophysiological mechanism responsible for sore pressures incidence did not seem to be simply linked to mechanical factors, such as pronation; it appeared instead to be related to an imbalance of multiple mechanisms (Coleman et al., 2013). The performed propensity score analysis tends to confirm this assumption, showing no protective factor of pronation in relation to the incidence of pressure injury.

Our study presented some limitations. Firstly, it was a monocentric observational retrospective study, with a relatively small series of patients. Secondly, due to the emergency situation caused by the pandemic, it was not possible to obtain a control group; nevertheless, the evidence that major complications directly related to pronation had almost disappeared, while minor complications (not directly related to pronation) incidence persisted, can be intended as “indirect control", suggesting that the presence of a specific team for pronation can actually reduce the rate of major complications related to this gesture. Thirdly, due to the retrospective nature of the study, only pressure injuries were considered among the minor complications investigated; however, among minor complications, pressure ulcers were the most important and the most represented. It can be speculated that pronation played a protective role only in complications related to a direct gesture.

## CONCLUSION

Compared to the current literature, the presence of the IPT seems to ameliorate the pronation gesture in ICU critical patients affected by COVID-19, resulting in a very low rate of major complications directly related to this technical gesture. The persistent high incidence of minor complications appeared to be a consequence of disease severity, independently from pronation itself, supporting the hypothesis that pressure injuries are induced by complex pathogenetic mechanism, probably related to multiple mechanisms and not frankly influenced by pronation.

## Data Availability

All data are available under specific request

## ACKNOWLEDGMENTS

We would like to thank Carlo Duca, for all the precious good advices.

